# Novel SARS-CoV-2 spike variant identified through viral genome sequencing of the pediatric Washington D.C. COVID-19 outbreak

**DOI:** 10.1101/2021.02.08.21251344

**Authors:** Jonathan LoTempio, Erik Billings, Kyah Draper, Christal Ralph, Mahdi Moshgriz, Nhat Duong, Jennifer Dien Bard, Xiaowu Gai, David Wessel, Roberta L. DeBiasi, Joseph M. Campos, Eric Vilain, Meghan Delaney, Drew G. Michael

**Affiliations:** Children’s National Hospital, Center for Genetic Medicine Research; George Washington University School of Medicine and Health Sciences, Department of Genomics and Precision Medicine; Children’s National Hospital Division of Pathology and Laboratory Medicine Department; Children’s Hospital of Los Angeles, Department of Pathology and Laboratory Medicine; Keck School of Medicine, University of Southern California; Children’s National Hospital, Division of Cardiology; Children’s National Hospital, Division of Critical Care Medicine; George Washington University School of Medicine and Health Sciences, Department of Pediatrics; George Washington University School of Medicine and Health Sciences, Department of Microbiology, Immunology and Tropical Medicine; George Washington University School of Medicine and Health Sciences, Department of Pathology; Children’s National Hospital, Division of Pediatric Infectious Diseases

**Author notes:** Address correspondence to Drew G. Michael.

## Abstract

The SARS-CoV-2 virus has emerged as a global pandemic, severely impacting everyday life. Significant resources have been dedicated towards profiling the viral genome in the adult population. We present an analysis of viral genomes acquired from pediatric patients presenting to Children’s National Hospital in Washington D.C, including 24 with primary SARS CoV2 infection and 3 with Multisystem Inflammatory Syndrome in Children (MIS-C) undergoing treatment at our facility. Viral genome analysis using next generation sequencing indicated that approximately 81% of the analyzed strains were of the GH clade, 7% of the cases belonged to the GR clade, and 12% of the cases belonged to S, V, or G clades. One sample, acquired from a neonatal patient, presented with the highest viral RNA load of all patients evaluated at our center. Viral sequencing of this sample identified a SARS-CoV-2 spike variant, S:N679S. Analysis of data deposited in the GISAID global database of viral sequences shows the S:N679S variant is present in eight other sequenced samples within the US mid-Atlantic region. The similarity of the regional sequences suggests transmission and persistence of the SARS-CoV-2 variant within the Capitol region, raising the importance of increasing the frequency of SARS-CoV-2 genomic surveillance.

**IMPORTANCE:** A variant in the SARS-CoV-2 spike protein was identified in a febrile neonate who was hospitalized with COVID-19. This patient exhibited the highest viral RNA load of any COVID-19 patient tested at our center. Viral sequencing identified a spike protein variant, S:N679S, which is proximal to the cleavage site at residue 681. The SARS-CoV-2 surface spike is a protein trimer (three subunits) which serves as the key target for antibody therapies and vaccine development. Study of viral sequences from the GISAID database revealed eight related sequences from the US mid-Atlantic region. The identification of this variant in a very young patient, its critical location in the spike polyprotein, and the evidence that it has been detected in other patients in our region underscores the need for increased viral sequencing to monitor variant prevalence and emergence, which may have a direct impact on recommended public health measures and vaccination strategies.

## INTRODUCTION

SARS-CoV-2, a positive-sense single-stranded RNA virus, is the causative agent of the ongoing COVID-19 pandemic (1–3). Reports in early 2020 suggested that children were spared the harshest manifestations of disease, with the majority of patients reported as asymptomatic (4). The first wave of outbreaks across Europe and the Americas demonstrated that this was not the case (5, 6), and in addition to the classical array of COVID-19 symptomatology, children were shown to be susceptible to a novel disease presentation, Multi-system Inflammatory Syndrome in Children (MIS-C) (7, 8).

Children’s National Hospital (CNH) in Washington, D.C., has evaluated and treated large numbers of pediatric and young adult patients with both primary SARS-CoV2 infection as well as MIS-C. The volume of patients seeking care has mirrored trends seen on the US East Coast, with an early spring 2020 peak, summer plateau, and an ongoing wave of infections which began in late November. As of January 12th, 2021, at least 821 Washingtonians have died of COVID-19, representing over 0.1% of the District of Columbia (D.C.) population (9), although none of these deaths have occurred in children at CNH, despite hundreds requiring hospitalization. Accordingly, our institution has allocated significant resources to understand the multi-factorial nature of SARS-CoV-2 primary infection and MIS-C.

The diagnosis of suspected COVID-19 patients at CNH, which includes D.C., Maryland, Virginia, West Virginia, and Delaware in its hospital catchment area, is routinely performed using semi-quantitative RT-PCR commercial platforms with EUA approved tests designed to assess multiple loci within the SARS-CoV-2 genome. When a test returns as positive, that patient enters treatment following a protocol designed to optimize care and resources. For selected samples, virus samples underwent viral sequencing to characterize the nature of the viral strains in our region.

Analysis of one viral isolate from a neonatal patient with a high viral RNA load resulted in the identification of a novel spike protein variant representing the earliest known sample of an emergent SARS-CoV-2 lineage in the US mid-Atlantic region. As the vaccine rollout for SARS-CoV-2 continues, the identification of this variant in the viral spike protein highlights the need for consistent, comprehensive molecular monitoring, transparency in reporting, and rapid response at all levels of healthcare and pandemic response systems.

## RESULTS

### Genomic profiling of pediatric COVID-19 SARS-CoV-2 virus in the Washington D.C. area identifies the GH clade as the predominant local viral clade

Seventy-six samples from COVID-19 patients presenting at CNH were selected for viral sequencing. Of these, twenty-seven samples met genetic sequencing coverage criteria for further analysis (>1,000 mean coverage), allowing for consensus sequences to be built and used for phylogenetic and variant analysis.

Viral sequences were analyzed via multiple sequence alignment (MSA) using ClustalW alongside representative samples from each viral clade as annotated in the Global Initiative on Sharing All Influenza Data (GISAID) repository and summarized in ***S1***. The phylogenetic relationship between our viral samples ***Fig. 1A***, allows for comparison to samples deposited from other laboratories in the D.C. area ***Fig. 1B***. These data indicate that the pediatric COVID-19 outbreak within the D.C. area followed trends observed in the adult population, as established by the clades of viruses deposited in the Global Initiative on Sharing All Influenza Data (GISAID) database and annotated for our geographic locale (***Fig. 1B*)**. This finding supports the hypothesis that the viral strains propagating in adults are similar to those in children.

**Fig 1.**
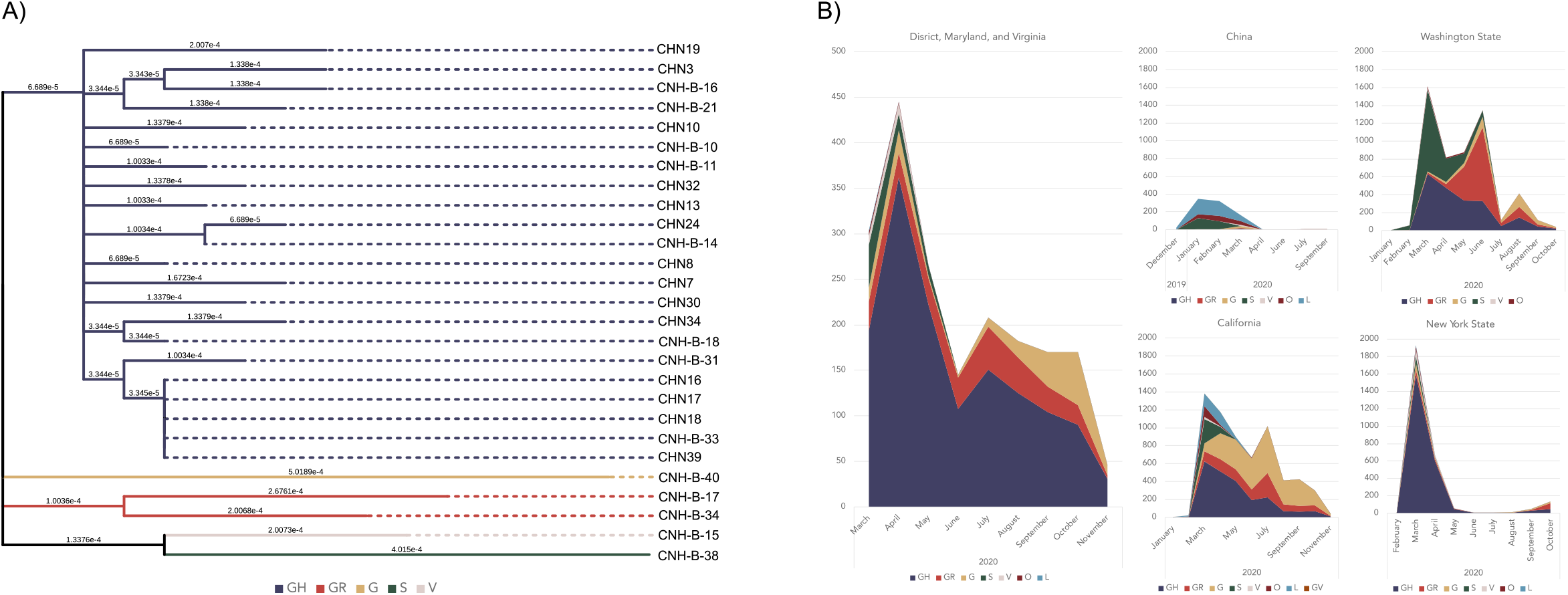
A) Maximum-likelihood phylogenetic tree of 27 SARS-CoV-2 positive patients. This tree was constructed from a ClustalW multiple sequence alignment and visualized with the Interactive Tree of Life GUI. Solid branches represent divergence in substitutions per nucleotide, while dashed lines are added for ease of interpretation. Colors represent the GISAID clade to which each sample belongs. **B) Time series plot of the GISAID clades of GISAID submissions over time in select cities**. Sequence metadata were accessed from GISAID to establish coarse differences in viral population diversity over time.

The D.C. outbreak mirrored the New York State (NYS) outbreak in terms of early predominance of sequences assigned to the GH clade. Genetic differences in outbreaks over time can be seen when examining key US West Coast outbreaks, as well as the early Chinese outbreak, where different clades of the virus reached equilibrium as the pandemic progressed (***Fig. 1B***). Analysis of the distribution of deposited viral sequences over time allows for the coarse analysis of predominant strains in each area. In the D.C. area, along with NYS, Washington State, and California. The ratio of clades present in each location changes with an increase in deposited viral sequences, demonstrating the evolving and dynamic nature of the SARS-CoV-2 variant profile across the pandemic.

To assess the possible geographic origin of viral strains propagating locally, we utilized the Phylogenetic Assignment of Named Global Outbreak LINeages, or PANGOLIN tool. PANGOLIN leverages data deposited at GISAID to assign lineages and find near global genetic neighbors based on their internal variant-based lineage assignment, which allows for sequence level analysis of global relationships. The countries which had the highest number of sequence submissions with the same lineages as our 27 samples were the US, United Kingdom (UK), and France, with minor relationships to sequences deposited from Portugal, Australia, Israel, and New Zealand (***Fig. 2***).

**Fig 2.**
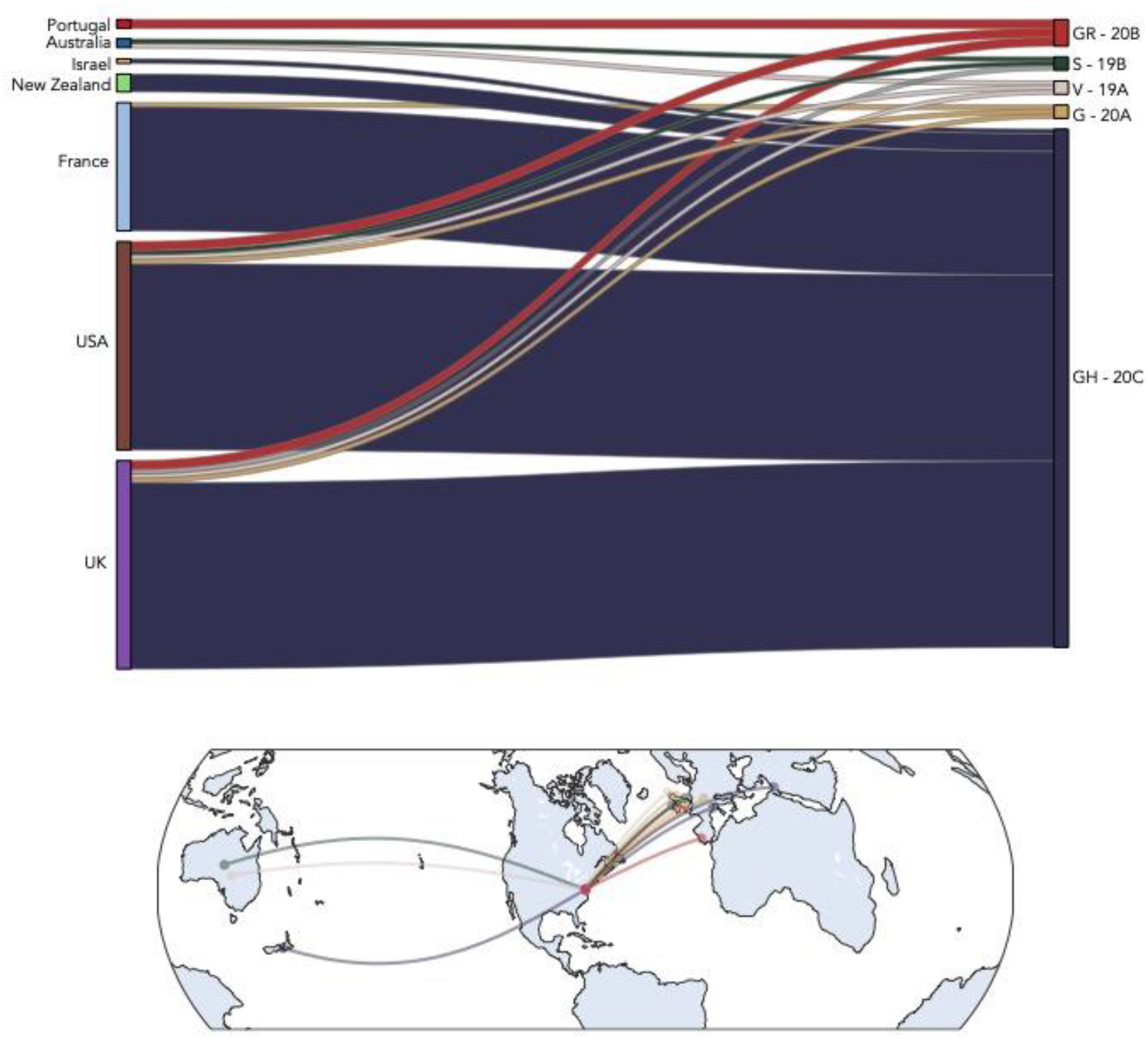
Global network of SARS-CoV-2. Representation of the locations where the highest number of sequences assigned to a given PANGOLIN lineage can be found in relation to DC sequences

The results point toward a European origin for the virus propagating in the US Capitol region pediatric population. The observation of the US and UK as major locations of genetic similarity is consistent with domestic population flux across US state borders, which remain porous, as well as international population flux, which was restricted with China on January 31, 2020, but unrestricted with Europe generally until March 11, 2020 (10, 11). Considering this, the incomplete nature of early travel restrictions does not appear to have had a protective effect on the US population. The UK is an outsized-GISAID contributor, responsible for sharing over 104,000 of the 263,000 high-quality, complete genomes in GISAID as of January 12, contributing to its weight in comparison to the samples sequenced in D.C. The third highest genetic contributor, France, was likely an early contributor to our outbreak in the D.C. area as determined by the presence of variants specific to the GH lineage samples found both in the Washington Metro region and first identified in France in February 2020 (DeBiasi et al, unpublished data under review). One limitation of the use of the PANGOLIN tool is that, since it is tied to the ever-expanding GISAID resource, relationships over time may change based on newly-deposited viral sequences.

### Viral variant profile of three pediatric MIS-C patients highlights the complexity of COVID-19 genotype-phenotype associations

To attempt correlation of SARS-CoV-2 variants with phenotypic outcomes, we identified consensus variants within the viral genome and overlaid the distribution of viral genetic variants with patient MIS-C clinical status. These analyses identified multiple variants present within each viral sample. Of note, 92% (25/27) of the viral genomes within the sampled cases contained the S:D614G spike variant associated with increased viral transmissibility (12).

Interestingly, five patients were identified with identical viral variant profiles. Four of these patients presented with primary COVID-19 disease and one presented with MIS-C. These samples are distinguished from other sequences assigned to the GH clade by the presence of variants in both the nucleoprotein (N:S193I) and non-structural protein 2 (NSP2:T371I), (***Fig. 3***).

**Fig 3.**
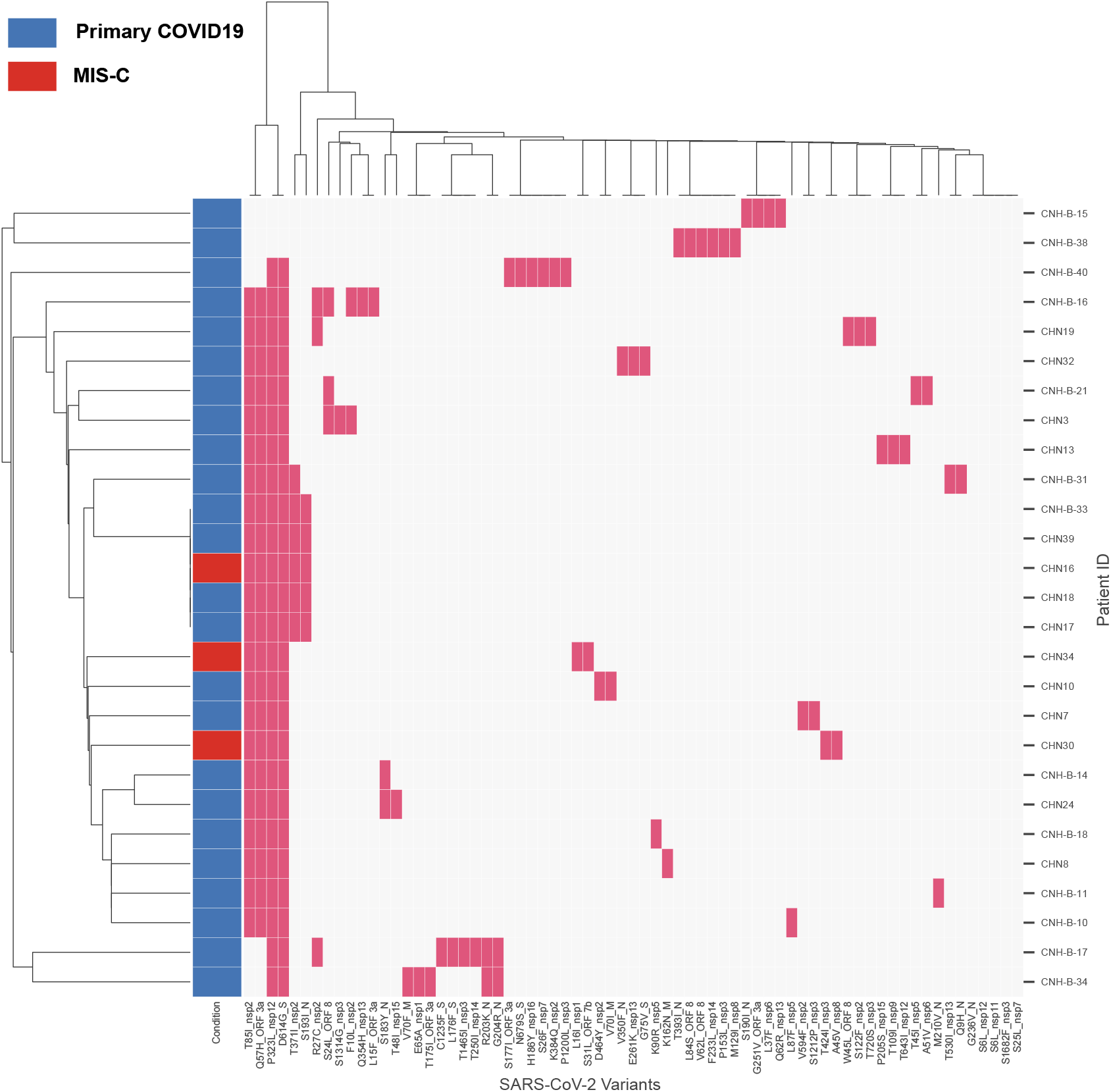
Clustering of detected SARS-CoV-2 viral genome variants with disease outcome. Bi-clustering of the binary variant matrix of SARS reveals a diverse cluster of variants and five patients with identical viral genomic profiles. The condition of each patient is depicted indicated primary COVID-19 (blue) and MIS-C (red). Variants identified across the cohort were aggregated and a binary matrix was generated for each patient and variant. Red colored boxes indicate the detection of a variant within that case.

In the GISAID database, 1198 sequences have N:S193I, while 277 have NSP2:T371I -only seven have both (GISAID accessions: EPI_ISL_516718, EPI_ISL_683413, EPI_ISL_745300, EPI_ISL_676627, EPI_ISL_683422, EPI_ISL_710051, EPI_ISL_683418). Six of those seven were generated from samples originating in Texas in late July, while the final sequence was generated from a sample originating in Virginia collected on April 30, 2020.

All patients presenting with MIS-C shared two GH clade-specific variants (NSP2:T85I and ORF3a:Q57H) with other samples from patients who presented with primary COVID-19. One, which was identical to other previously-discussed samples, had nucleocapsid variants N:S193I and NSP2:T371I. The second had the additional variant NSP1:L16I, while the third had NSP3:T424I and NSP8:A45V.

The observation of five identical viral genomic profiles with different disease outcomes, and two clinically similar MIS-C cases with different viral genotypes highlights the difficulties inherent in forming viral genotype-human phenotype correlations in this disease (***Fig. 3***). This observation points toward the importance of additional variables, such as initial viral dosage, environmental factors, and host genetic predispositions as key elements of COVID-19 progression and possibly underlying MIS-C presentation.

### High viral RNA load on presentation in neonatal patient leads to identification of novel spike protein S:N679S variant

In early September 2020, a febrile neonatal patient was hospitalized at CNH with COVID-19 symptoms. SARS-CoV-2 RT-PCR testing detected a high diagnostic viral load as measured by the polymerase chain reaction (PCR) cycle threshold (Ct) of 6.45 using an assay which targets ORF (Ct 6.4) and spike (Ct 6.5) (Simplexa COVID-19 Direct, Diasorin, CA, USA). In comparison, we observed a median Ct of 22.1 for both genes during 499 previous positive tests. The observed Ct for this patient represented an approximate 51,418-fold increase in viral load compared to the median presenting viral load in previously seen patients. Supplemental testing of this patient’s sample on a different platform yielded a Ct of 6.5 (Allplex^™^ 2019-nCoV Assay, Seegene, S Korea) confirming the extremely high viral load. As this patient represented an outlier in viral load, we selected this sample for NGS profiling of the SARS-CoV-2 genome.

Short read sequencing analysis of the SARS-CoV-2 genome identified a novel variant which resulted in the nonsynonymous amino acid substitution S:N679S, which is responsible for viral entry. The presence of >9,000 individual reads was strong support for a spike protein variant, rather than a sequencing error or artifact of PCR (***Fig. 4B***). In order to confirm with an orthogonal sequencing technology, the sample was sent for confirmation of the variant by Sanger sequencing, which confirmed the presence of the S:N679S variant (***Fig. 4C***). The Sanger data analysis also confirmed the presence of the S:D614G variant in the viral genome, which is present in the majority of global samples due in part to its hypothesized greater infective potential (12). This is significant, as association of the S:N679S with S:D614G may contribute to the persistence of the S:N679S variant.

**Fig 4.**
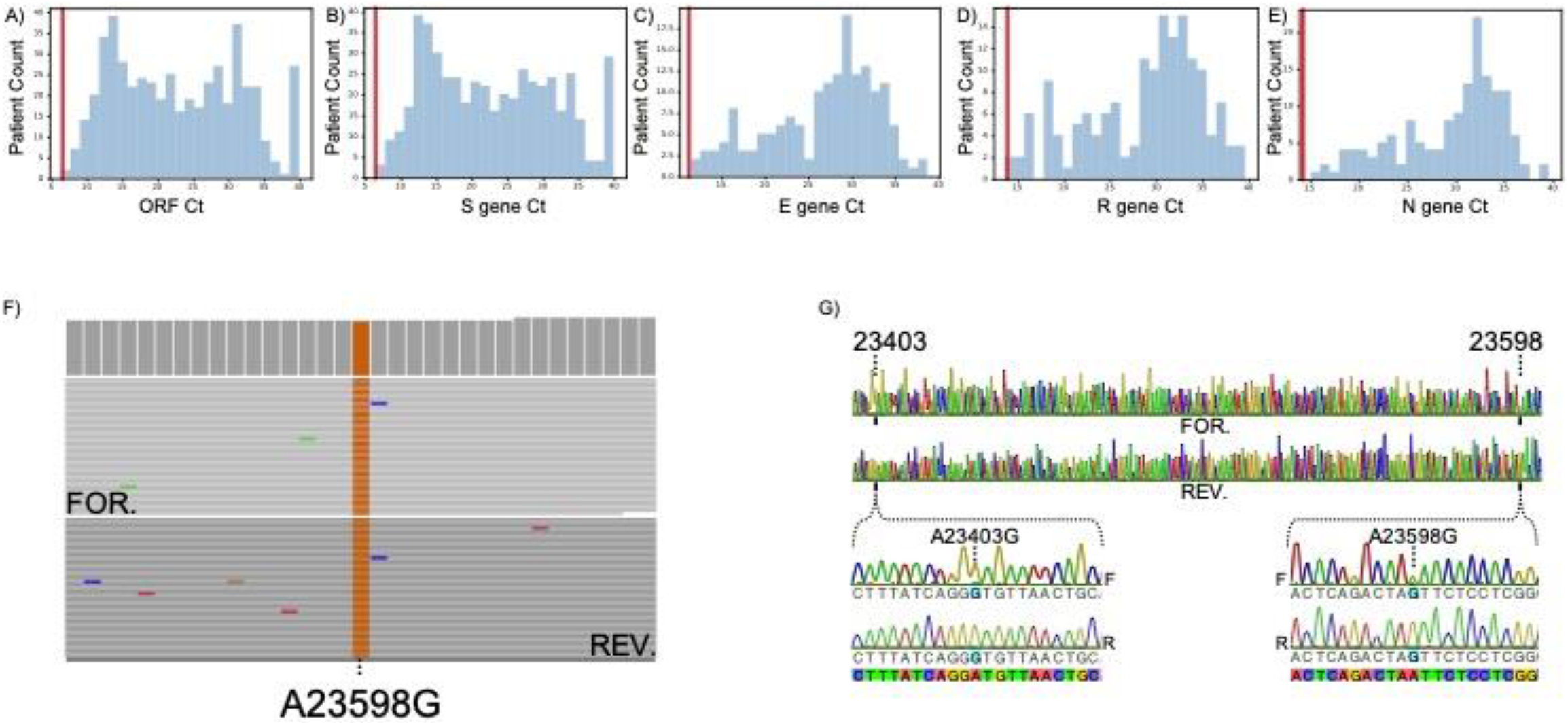
A-E) Histograms of viral PCR test cycle threshold values. Vertical red line indicates the detected RT-PCR of the S:N679S case. **A)** Diasorin ORF Ct distribution **B)** Diasorin S gene Ct distribution. **C)** Seegene RT-PCR E gene Ct distribution. **D)** Seegeen R gene Ct distribution. **E)** Seegene N gene Ct distribution. **F)** Short-read pileup at the coding region for spike protein variant of interest, S:N679S. The read pileup shows strong homozygous signal (>9,000x coverage) for a non-synonymous variant in the viral spike protein. **G)** Sanger sequence confirmation of variant of interest, S:N679S. Chromatogram showing a contiguous span containing the previously characterized D614G variant and linkage to the S:N679S variant of interest.

To assess where the S:N679S variant is present in the community, we queried the GISAID database. At the time of the initial query, the GISAID database contained six high-quality complete genomes containing the S:N679S variant. All six of these genomes were deposited by labs in Maryland and Virginia. In mid-December, 2020 re-query identified an additional four samples from Australia and Japan, with a third re-query revealing a sequence from Brazil. Finally, a fourth query on January 12, 2021 revealed two more high-quality sequences from Delaware. Phylogenetic analysis (***Fig. 5A)*** showed that this novel spike variant had emerged in two distinct viral clades which are geographically and genetically independent of each other. Variant profiles and lineage assignments suggest the singleton samples from Brazil and Japan, and the samples from Australia, while all in clade GR, are genetically distinct in lineages B.1.1.33, B.1.1.284, and B.1.1.162, respectively. Conversely, all samples collected in the US were assigned to the lineage B.1.189 by PANGOLIN, further supporting their similarity within the large G clade.

**Fig 5.**
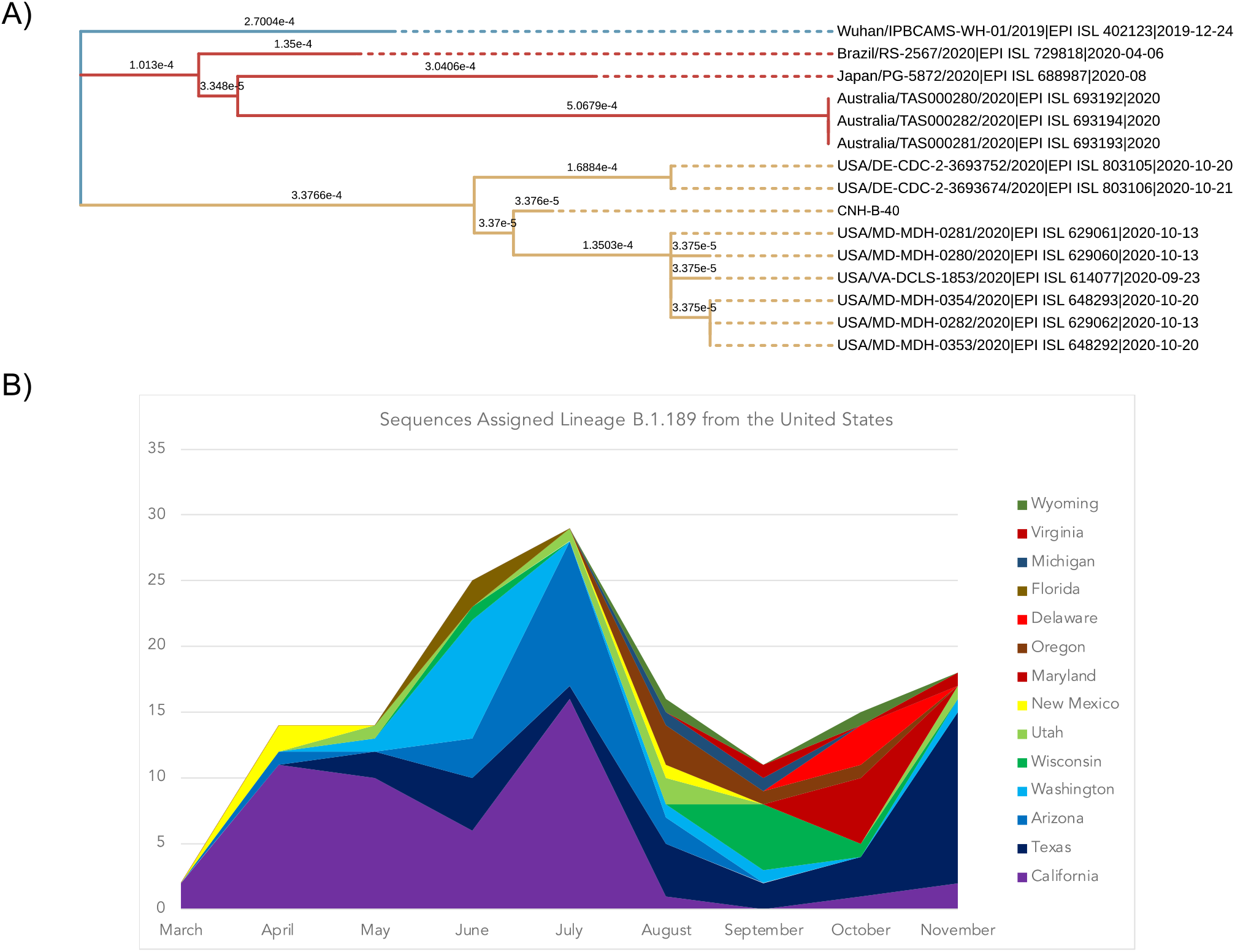
A) Maximum likelihood phylogenetic tree of all high-quality sequences in GISAID with variant S:N679S in the spike protein. A non-reference sample from Wuhan collected in December 2019 was used as the outgroup in this tree to draw contrast between clades. Clade G in yellow shows the high-quality sequences in lineage B.1.189 where spike variant S:N679S has emerged. Clade GR in yellow shows three separate evolutionary events resulting in spike variant S:N679S. **B) Time series plot of the count, by location, of sequences in GISAID belonging to PANGOLIN lineage B.1.189**. Sequence metadata were accessed from GISAID to establish coarse differences in viral population diversity over time.

To probe the similarity between the reported samples of US origin, we constructed a maximum-likelihood (ML) phylogeny with international samples for context ***(Fig. 5A)***. The tree topology among US mid-Atlantic regional samples is consistent with time-of-sampling metadata, which is available for all US samples. The patient treated at CNH, who exists in a branch distinct from the other six samples from the D.C. area, was sampled on 17 days before the next earliest sample on September 23rd. Both samples from Maryland collected on October 20, as well as one from October 13 are genetically identical with branch length zero. It is remarkable that samples collected on the 20th and 21st of October in Delaware share a common ancestor predating the samples found locally (including our sample from September 7, 2020). This suggests both the persistence of the variant, and earlier emergence than was previously appreciated with samples from the District of Columbia, Maryland, and Virginia.

### Spike protein residue 679 is proximal to the S1/S2 cleavage site and may impact function

We queried GISAID for all amino acid substitutions at spike polyprotein position 679, the results of which are presented in ***S2_a***. The most common variants observed so far include substitution with lysine or deletion, both of which represent 703 of 731 variants described in all human sequences in GISAID. Substitution with serine, the variant observed at CNH, represented 16 human sequences as of January 12, 2021. Five substitutions to histidine, two to isoleucine, two to tyrosine, and one to threonine were also observed. There are over 370,000 sequences containing the wild-type asparagine.

The relatively high number of lysine variants can be partially attributed to codon wobble -both have tRNA three letter codes AAX, with asparagine coded by AAU and AAC, while lysine is coded by AAA and AAG. The serine, threonine, and tyrosine require transition or transversion at the second position of the codon, while transversion of the first codon position is required to result in histidine. A multifasta (***S2_b***) with each of these amino acid substitutions, and a deletion of the residue at 679 was constructed and analyzed with the ProP tool for predicting cleavage sites (13). With the exception of the S:N679Y variant, each substitution scored higher than the wild type threshold of 0.62. Higher ProP scores indicate a higher likelihood the site will function as a site for cleavage. This suggests our observed S:N679S substitution at this residue does not preclude the variant protein from activation. A robust and rapid pipeline for functional classification of SARS-CoV-2 variants will be an important public health tool as the global community continues to navigate the COVID-19 pandemic.

In 2011, it was shown that SARS-CoV spike protein is cleaved and activated by human airway trypsin-like protease (HAT)(14). Furthermore, it has been suggested that the insertion of the RRAR amino acid sequence at the S1/S2 cleavage site in SARS-CoV-2, relative to SARS-CoV, represents a gain of function as a target of human peptidase furin, which is known to cleave basic amino acids at the motif R–X–[R/K]–R. This motif is indeed observed between S1 and S2 spike subunits in SARS-CoV-2. *In silico* and benchtop experimental evidence has been presented for SARS-CoV-2 variants with and without this motif showing SARS-CoV-2 variants lacking the RRAR motif are unable to fuse with HEK293 cells in the absence of trypsin or HAT, while the wild type SARS-CoV-2 spike can undergo cellular fusion without HAT or trypsin (15).

This is strong evidence for a cell fusion gain of function through the RRAR protein motif – but not for an increase in virulence or infectivity. Indeed, SARS-CoV-2 variants where the furin cleavage site has been deleted show increased infectivity (16, 17). Whether preservation of the furin cleavage site and its gain of function for cell fusion relative to SARS-CoV is outweighed by the increase in infectivity in its absence (as in SARS-CoV-2) should be functionally assessed in cell and animal experiments.

### Analysis of global SARS-CoV-2 data provides evidence for mid-Atlantic circulation of the spike variant S:N679S

As of January 12, 2021, there are 161 sequences in GISAID which are annotated as B.1.189 (including low quality sequences) which represent the nearest genetic neighbors of our sample of interest. Of these sequences, 149 are from the US, nine are from Mexico, two are from Canada, and one is from Japan. The earliest available sequences were from samples collected in March in California and Mexico City. Two samples from Manitoba, Canada, were collected in May, and the single sample from Japan was collected in July. Examination of the temporal and geographic distribution of this lineage further suggests circulation in North America, from coast to coast, and across the Mexican-American, and Canadian-American borders ***(Fig. 5 B)***. Variant analysis with NextClade (18) shows only two silent mutations present inside and outside the D.C. area -A20268G and C4582T -suggesting the possibility of many missing representatives of the lineage.

The S:N679S-containing sample detected at CNH has silent mutation C2710T, which is absent in all other local samples. All D.C. area samples contain two missense mutations, ORF1a:A2994V and ORF1a:P3359S, as well as silent mutation T11408C. The difference between these samples suggests there is likely an earlier case which has the additional variants common to all seven samples including missense mutations ORF1a:K564Q, ORF1a:P2018L, ORF1a:S3885F, ORF1b:H2583Y, and ORF3a:S177I, as well as silent mutations C4276T, T6160C, C16293T, C16887T, C26222T. This predicted most recent common ancestor, likely appeared in summer 2020. Additional sequencing of archived samples will be required to assess this hypothesis.

When considering that there are two high-quality sequences in the database are from samples acquired in Delaware, the timeline can be revised to include longer circulation and wider spread. Those samples contain the coding variants ORF1a:S3885F, ORF1b:P314L, ORF1b:H2583Y, ORF3a:S177I, S:D614G, S:N679S, which are common to the samples from the D.C. area. One sample, EPI_ISL_803105 collected on October 20, also has ORF1a:K564Q which is common to the D.C. region samples, while the other, EPI_ISL_803106 collected one day later, does not have that variant which possibly represents a loss of the variant through back mutation to the reference nucleotide, A, at position 1955. These sequences also contain further coding variants ORF10:Q29* and ORF1a:C1114F, but lack all other ORF1a variants.

Taken together, these data support the hypothesis wherein a SARS-CoV-2 virus containing the variant S:N679S is circulating within the US mid-Atlantic region. Based on these analyses, the ancestral state is hypothesized to include the following variants: ORF1a:K564Q, ORF1a:S3885F, ORF1b:P314L, ORF1b:H2583Y, ORF3a:S177I, S:D614G, S:N679S. All other variants are lineage specific. All variant profiles of these sequences can be found in S1.

### Low sequencing rates in the US hinder surveillance of emerging viral variation

As of January 12, the US has passed 25 million reported cases and 433,000 reported deaths; the range of excess deaths in the US reported by the CDC and attributed to COVID-19 was given a lower bound of 346,750 and an upper bound of 469,831 (19). Concurrently, approximately 71,000 sequences deposited to GISAID are from the US, representing ∼0.3% of confirmed cases. We leverage data from Australia (59.6% of confirmed cases, or 17,081/28,650 cases sequenced), Japan (3.3% or 9,885/298,000 cases), and Brazil (0.02% or 2,102/8.2m cases). This differs from the UK, the world’s largest COVID-19 viral genome contributor with 157,626 sequences deposited from 3.16 million cases (∼4.99%). All data are based on GISAID submissions and the Johns Hopkins’ interactive dashboard (20).

Our analysis of the S:N679S containing viral genome from sample CNH-B-40 began in early December; other local high-quality sequences with the variant had been deposited just weeks prior. In the intervening four weeks, the sequences from Australia, Japan, and Brazil were deposited, along with another low-quality sequence from Delaware in the US. This brings the total number of high-quality sequences from human samples in GISAID with S:N679S to twelve, including CNH-B-40 which is yet to be assigned a GISAID accession number.

The highly-related samples with S:N679S were all collected over a large, interconnected 4-state/district geographic area in a relatively short 7-week timeframe with no established person-to-person transmission network. Given the high likelihood of community circulation of this variant of SARS-CoV-2, it is possible that additional cases of this variant strain are present in the mid-Atlantic region but have yet to be detected due to low rates of viral sequencing and deposition to GISAID.

As of January 12, Delaware has approximately 67,000 confirmed cases and 345 sequences deposited (0.5%), Maryland has 315,000 confirmed cases and 802 sequences deposited (0.25%). D.C. has 33,000 confirmed cases and 130 sequences deposited (plus our 27) (0.4%, 0.47% combined), and Virginia has 413,000 confirmed cases and 1,685 sequences deposited (0.4%). Taken together, in states from this region, there are at least 828,000 cases, but only 2,962 sequences. This represents a sequencing rate of 0.36%, just over the national average of 0.3%, but considerably lower than the 4.99% of samples sequenced in the UK, where a critical new variant was first identified (21, 22).

## DISCUSSION

A key goal of precision medicine is to target care to those who need it most. An ideal COVID-19 diagnostic and triage algorithm would integrate features such as viral genomic profile, host innate errors in immunity and viral load over the course of infection to inform care. Toward this end, we established systems to profile the SARS-CoV-2 genome and link viral genotype to pediatric disease outcomes. This report contains the initial analysis of twenty-seven patients and highlights the complexity of generating effective viral genotype-human phenotype correlations. Given the complex, multifactorial nature of COVID-19, larger pediatric studies which link to phenotypic outcomes will be required.

We note the parallel between the emergence of antibiotic resistance traits in bacteria (23, 24) and viral evolution. Physical and social distancing measures have presented a transmission and genetic bottleneck to SARS-CoV-2, with data supporting a different mix of virus clades in each successive wave. The wide-scale development and deployment of vaccines for SARS-CoV-2 will present further bottlenecks to the virus. These measures, while demonstrably effective, have the potential to drive the evolution of strains capable of bypassing defenses. The emergence of the D614G variant and the increased infectivity of the UK variant strain B.1.1.7 / UK VUI 202012/01 strain highlight the potential for stochastic genomic variants to generate vaccine escaping viral strains and to exhibit altered SARS-CoV-2 fitness (12, 22).

At present, there is high-quality, genomic evidence which supports the biological tolerance or persistence of the S:N679S variant in at least two distinct clades. This includes the variant S:N679S in the GR clade found in Brazil, Japan, and Australia, and the variant found in G clade in September and October of 2020 in the US mid-Atlantic region. While the sequences from Brazil, Japan, and Australia have been assigned the GR clade, they do not belong to the same PANGOLIN lineage due to variation in their genomes, which we can see recapitulated in the tree topology of **Fig. 5a**. This is strong evidence for four different evolutionary events which are tolerated in GR and G SARS-CoV-2 clades.

The UK presents a strong case for widespread use of high throughput sequencing technologies and rapid data sharing so that discoveries like these are more rapidly made, confirmed, and acted upon. Under current funding levels, the US CDC plans to fund the sequencing and release of at least 6,000 viruses per week as reported on January 4, 2021 (25, 26). This represents approximately 0.4% of daily cases based on current daily caseloads in excess of 200,000. This is still well below the UK’s level of sequencing (4.99%), but is a commendable step in the right direction, assuming that samples that are sequenced will be representative of the population, allowing for surveillance. Inclusion of data on disease severity, patient age and ethnicity, co-morbidities, and other relevant contextual data will help researchers ascertain the generalizability of sequences in hand, or adjust for confounding factors as needed. Further funds should be allocated to promote a collaborative effort to sequence biobanked samples in an effort to understand viral evolution and transmission paths.

Our analyses identified the S:N679S variant within a neonatal patient with a high observed viral load at presentation. This single case observation currently represents insufficient evidence to propose a causal relationship between the S:N679S variant and increased viral loads or presentation at a very young age. Analysis of the GISAID data for pediatric enrichment was not possible due to the lack of patient metadata for records with this variant. The observation that this variant strain of SARS-CoV-2 is currently undergoing community transmission in the US mid-Atlantic area warrants continuous and rigorous monitoring. The SARS-CoV-2 spike protein not only moderates viral infectivity and cellular uptake, but is also a target for vaccine and monoclonal antibody therapeutic development. While vaccines are designed to elicit a polyclonal immune response, the primary target of the vaccine response is currently a monogenic antigen, and monoclonal antibody cocktails like those produced earlier this year run the risk of selection for spike variants that escape those treatments (27).

In conclusion, a new protein coding spike variant, S:N679S, has been discovered in a clinically-unique patient in Washington, D.C. Analysis of global data shows that presence of the variant was tolerated by SARS-CoV-2 in at least four different viral lineages, one of which shows evidence of transmission of the variant to recipients in the US mid-Atlantic region. One of these lineages presented within the CNH catchment area, showing viral sequence similarity to our case. Variation in emerging infectious disease is expected, but this finding underscores the need for larger-scale whole viral genome monitoring to ensure that responses can be quickly adapted in the face of viral evolution. Until then, variants will continue to circulate undetected, hindering the global response to SARS-CoV-2. Progress toward the defeat of the COVID-19 pandemic will require effective application of public health measures and wide-scale vaccination alongside careful monitoring for emergence of viral escape variants.

## MATERIALS AND METHODS

### Initial COVID-19 Screening

Initial COVID-19 screening was performed at CNH using either the Diasorin Liaison® MDX-Simplexa^™^ COVID-19 Direct real-time PCR assay or the Seegene Allplex^™^ 2019-nCoV real-time PCR assay. Both assays are qualitative and can be performed using nasopharyngeal swab, oropharyngeal swabs, or saliva collections. The Diasorin Molecular Simplexa COVID-19 assay allows for direct amplification of the ORF1ab region and the S gene (Spike glycoprotein) present in the viral sample. The Seegene Allplex COVID-19 assay requires viral RNA extraction prior to amplification and uses the Bio-Rad CFX96 real-time PCR thermocycler for detection. This assay uses primers designed to detect the presence of the E gene (an envelope protein), RdRP (RNA-dependent RNA polymerase), and the N gene (a nucleoprotein).

### RNA extraction and cDNA generation

Viral RNA was extracted with the Qiagen EZ1® DSP Virus Kit, a magnetic bead-based kit. The concentration of the viral RNA was assessed using the NanoDrop One. To perform Sanger confirmation on targeted variants, the viral RNA underwent reverse transcriptase cDNA generation using the High Capacity cDNA Reverse Transcription kit manufactured by Applied Biosystems. The RT master-mix was prepared according to manufacturer recommendations excluding the 10X RT Random Primers, as we had designed our own primers based on the sequencing data. To avoid potential primer bias, cDNA was synthesized using three different initiation sites in the viral genome

### Viral sequencing

Sequence data was generated on the Illumina MiSeq platform as a part of a collaborative effort between CNH and Children’s Hospital of Los Angeles with institutional review board approval. Following previously described methodology (28), the CleanPlex SARS-CoV-2 NGS Panel kit (Paragon Genomics, Inc., Hayward, CA) was used to produce cDNA and then amplify the SARS-CoV-2 genome in two multiplex PCR reactions containing 171 and 172 primer pairs each, producing amplicons ∼150bp in length, and covering nearly the entire viral genome once assembled (∼100bp missing on each end). Assembly was performed using the NovoAlign v4.02.00 algorithm with NC_045512 as the SARS-CoV-2 reference sequence.

### Sanger confirmation

At completion of reverse transcription, Sanger sequencing of selected variants identified by NGS was performed. We utilized the AccuPrime^™^ *Taq* DNA Polymerase System manufactured by Invitrogen^™^. A total of 10 PCR reactions containing different forward and reverse primer pairings were prepared to confirm the observed variants. Thermocycler conditions: denature 94°C 30s, anneal 55°C 30s, extend 68°C 60s, for 30 cycles. The final PCR products were run on a 2% E-Gel to assess for presence of bands and fragment sizes. The PCR products were Sanger sequenced by GeneWiz (Frederick, MD).

#### RT primers

23902r* TAAATTTGTTTGACTTGTGCAAAAACT

24527r CCTGTGATCAACCTATCAATTTGCACTTCAGC

24609r AAGATTAGCAGAAGCTCTGATTTCTG

#### PCR primers

23320_M13F* CATTACACCATGTTCTTTTGGTGGTGT

23727_M13R* TAGACACTGGTAGAATTTCTGTGGTAA

23381_M13F CAGGTTGCTGTTCTTTATCAGGGT

23902_M13R TAAATTTGTTTGACTTGTGCAAAAACT

23564_M13F GCAGGTATATGCGCT

24038_M13R GCCAGCATCTGCAAGTGTCAC

23902_M13F TTTTGCACAAGTCAAACAAATTTA

24570_M13R GTTGAGTCACATATGTCTGCAAAC

*used to generate the representative chromatogram shown in **Fig 4**. Primer sequences shown 5’ to 3’ for their respective strands. M13 sequencing tags have been removed for clarity.

### Data access

GISAID is presently the leader in viral sequence data sharing, having rapidly expanded their influenza data sharing capabilities to suit the COVID-19 pandemic (29). All data from samples outside of CNH were accessed from the GISAID database in accordance with their data sharing agreement (30). As of submission, sequences from CNH have not been assigned GISAID accession IDs, but will be available in that repository.

### Phylogenetics

Consensus sequences were generated from variant calls with bcf tools on the SARS-CoV-2 reference sequence from Wuhan, China (NC_045512) (31). MSA were generated from consensus sequences from CNH samples and from GISAID through ClustalW alignment on default settings (32). An ML phylogenetic tree was generated on the MSA with MEGA 7.0 (33, 34). All tree files were visualized with Interactive Tree of Life (35).

### Variant analysis

The NextClade and CoV-Glue tools were used for variant analysis (18, 36) and compared to clade, lineage, and global distribution of related sequences via the PANGOLIN tool. NextClade assesses variation by aligning a query sequence to the SARS-CoV-2 reference sequence by scanning 21-mers across the genome followed by a banded Smith-Waterman alignment with affine gap penalty, while CoV-GLUE leverages MAFFT (38) and RAxML (39), with detailed methods described in their preprint. PANGOLIN leverages MAFFT and IQ-TREE 2 (40), with methods previously described. All relevant files are available as ***S3***.

### Proteinase cleavage site prediction

A multifasta of S1/S2 cleavage site was uploaded to ProP1.0 for analysis and prediction of cleavage sites (41). Scores were exported and organized by of GISAID variants. ***S2***.

## Supporting information

Supplemental_files_1_through_4

## Data Availability

All data with GISAD access numbers are available at www.gisaid.org
All data without GISAID accession numbers are being submitted to GISAID or available from the authors in the interim.

## Acknowledgements

We acknowledge the critical efforts of Kristen Kocher, Melissa Andrew, and Miguel Almalvez who made viral transport media and sample collection kits during the shortage which occurred in spring and summer 2020.

We acknowledge Tara Workman, Hasani Malcolm, Farzana Siddiqui, Katy Draper, and Ashenafi Melkamu who ran SARS-CoV-2 PCR tests; Eric Freemen, William Suslovic, and Aszia Burrel who coordinated the clinical research program; Michael Evangelista and Joyce Granados who contributed from the microbiology department.

We acknowledge the GISAID consortium for their comprehensive work. We acknowledge the specific originating labs: Maryland Public Health Laboratory, Virginia Division of Consolidated Laboratory Services, Royal Hobart Hospital, Australia, Pathogen Genomics Center at the Japan National Institute of Infectious Disease, Labrotorio Central de Saude Publica do Estado do Rio Grande do Sul, and Respiratory Viruses Branch, US CDC. (***S4***)

Our funders included the Ikaria Fund, and the A. James Clark Distinguished Professorship, CNH and the Saban Research Institute at CHLA intramural support for COVID-19 Directed Research.

## SUPPLEMENT LEGEND

**S1: List of case IDs, clades, and disease status**

S1_Case_clade_and_disease_status_12_21_2020.csv

**S2_A: ProP scores**

S2_A_GISAID_Spike_residue_679_variant_scores_15_jan_2021.pdf

**S2_B: Amino acid substitution multifasta**

S2_B_679_aa_sub.fa

**S3_A: Coding and non-coding variants in sequences with S:N679S**

S3_A_B1189_N679S_variant_summary.pdf

**S3_B: Variants called via NextClade**

S3_B_NextClade_variants.xlsx

**S4: Acknowledgements from originating labs, GISAID**

S4_gisaid_hcov-19_acknowledgement_table_high_quality_spike_n679S.pdf

## Notes

### Competing Interest Statement

The authors have declared no competing interest.

### Author Declarations

Expedited IRB review at Children's National Hospital Title: Genomic Epidemiology and Pathogenesis of COVID-19 in Children. No.: Pro00015182

