## Supplemental_files_1_through_4 for "Novel SARS-CoV-2 spike variant identified through viral genome sequencing of the pediatric Washington D.C. COVID-19 outbreak": S2_A_GISAID_Spike_residue_679_variant_scores_15_jan_2021.pdf

| Supplemental 2 | AA | AA_3 | AA_1 | Count N679-sub | Complete | Complete and high coverage | ProP1.0 Score |
|---|---|---|---|---|---|---|---|
| Positive | Arginine | Arg | R | 0 | 0 | 0 | |
| | Histidine | His | H | 5 | 5 | 0 | 0.641 *ProP* |
| | Lysine | Lys | K | 325 | 323 | 294 | 0.648 *ProP* |
| Negative | Aspartic acid | Asp | D | 0 | 0 | 0 | |
| | Glutamic acid | Glu | E | 0 | 0 | 0 | |
| Polar | Serine | Ser | S | 16 | 16 | 13 | 0.651 *ProP* |
| | Threonine | Thr | T | 1 | 1 | 1 | 0.666 *ProP* |
| | Asparagine | Asn | N | wt | wt | wt | 0.620 *ProP* |
| | Glutamine | Gln | Q | 0 | 0 | 0 | |
| Special | Cystine | Cys | C | 0 | 0 | 0 | |
| | Glycine | Gly | G | 0 | 0 | 0 | |
| | Proline | Pro | P | 0 | 0 | 0 | |
| Hydrophobic | Alanine | Ala | A | 0 | 0 | 0 | |
| | Valine | Val | V | 0 | 0 | 0 | |
| | Isoleucine | Ile | I | 2 | 2 | 2 | 0.633 *ProP* |
| | Leucine | Leu | L | 0 | 0 | 0 | |
| | Methionine | Met | M | 0 | 0 | 0 | |
| | Phenylalanine | Phe | F | 0 | 0 | 0 | |
| | Tyrosine | Tyr | Y | 2 | 2 | 2 | 0.616 *ProP* |
| | Tryptophan | Trp | W | 0 | 0 | 0 | |
| | Deletion (*) | | | 378 | 374 | 315 | 0.680 *ProP* |
| | | | Total variants | 729 | 723 | 627 | |
| | | | Presumed wt | 370,761 | 370,094 | 268,419 | |
