## Supplemental_files_1_through_4 for "Novel SARS-CoV-2 spike variant identified through viral genome sequencing of the pediatric Washington D.C. COVID-19 outbreak": S3_A_B1189_N679S_variant_summary.pdf

### Supplemental 2A: B.1.189 variation summary

#### B.1.189 S:N679S Nt Substitutions

| | 241 | 1955 | 2710 | 3037 | 3606 | 4276 | 4582 | 5672 | 6160 | 6318 | 9246 | 10336 | 10340 | 11200 | 11408 |
|---|---|---|---|---|---|---|---|---|---|---|---|---|---|---|---|
| CNH-B-40 | C241T | A1955C | C2710T | C3037T | | C4276T | C4582T | | T6160C | C6318T | | | | | |
| USA/VA-DCLS-1853/2020\|EPI_ISL_614077\|2020-09-23 | C241T | A1955C | | C3037T | | C4276T | C4582T | C5672T | T6160C | C6318T | C9246T | C10336T | C10340T | | T11408C |
| USA/MD-MDH-0280/2020\|EPI_ISL_629060\|2020-10-13 | C241T | A1955C | | C3037T | | C4276T | C4582T | | T6160C | C6318T | C9246T | C10336T | C10340T | C11200T | T11408C |
| USA/MD-MDH-0281/2020\|EPI_ISL_629061\|2020-10-13 | C241T | A1955C | | C3037T | | C4276T | C4582T | | T6160C | C6318T | C9246T | C10336T | C10340T | | T11408C |
| USA/MD-MDH-0282/2020\|EPI_ISL_629062\|2020-10-13 | C241T | A1955C | | C3037T | | C4276T | C4582T | | T6160C | C6318T | C9246T | C10336T | C10340T | | T11408C |
| USA/MD-MDH-0353/2020\|EPI_ISL_648292\|2020-10-20 | C241T | A1955C | | C3037T | | C4276T | C4582T | | T6160C | C6318T | C9246T | C10336T | C10340T | | T11408C |
| USA/MD-MDH-0354/2020\|EPI_ISL_648293\|2020-10-20 | C241T | A1955C | | C3037T | | C4276T | C4582T | | T6160C | C6318T | C9246T | C10336T | C10340T | | T11408C |
| USA/DE-CDC-2-3693752/2020\|EPI_ISL_803105\|2020-10-20 | C241T | A1955C | | C3037T | G3606T | C4276T | C4582T | | T6160C | | | | | | |
| USA/DE-CDC-2-3693674/2020\|EPI_ISL_803106\|2020-10-21 | C241T | | | C3037T | G3606T | C4276T | C4582T | | T6160C | | | | | | |

| | 11919 | 14408 | 15756 | 15879 | 16293 | 16887 | 20268 | 21214 | 23403 | 23598 | 25642 | 25922 | 26222 | 28957 | 29642 | 29738 | 29868 | 29900 |
|---|---|---|---|---|---|---|---|---|---|---|---|---|---|---|---|---|---|---|
| CNH-B-40 | C11919T | C14408T | | | C16293T | C16887T | A20268G | C21214T | A23403G | A23598G | | G25922T | C26222T | | | C29738T | | |
| USA/VA-DCLS-1853/2020\|EPI_ISL_614077\|2020-09-23 | C11919T | C14408T | | | C16293T | C16887T | A20268G | C21214T | | A23598G | | G25922T | C26222T | | | C29738T | | |
| USA/MD-MDH-0280/2020\|EPI_ISL_629060\|2020-10-13 | C11919T | C14408T | | | C16293T | C16887T | A20268G | C21214T | | A23598G | | G25922T | C26222T | | | C29738T | G29868C | |
| USA/MD-MDH-0281/2020\|EPI_ISL_629061\|2020-10-13 | C11919T | C14408T | | | C16293T | C16887T | A20268G | C21214T | A23403G | | | G25922T | C26222T | | | C29738T | | A29900G |
| USA/MD-MDH-0282/2020\|EPI_ISL_629062\|2020-10-13 | C11919T | C14408T | | | C16293T | C16887T | A20268G | C21214T | A23403G | | C25642T | G25922T | C26222T | | | C29738T | | |
| USA/MD-MDH-0353/2020\|EPI_ISL_648292\|2020-10-20 | C11919T | C14408T | | | C16293T | C16887T | A20268G | C21214T | A23403G | | C25642T | G25922T | C26222T | | | C29738T | | |
| USA/MD-MDH-0354/2020\|EPI_ISL_648293\|2020-10-20 | C11919T | C14408T | | | C16293T | C16887T | A20268G | C21214T | A23403G | | C25642T | G25922T | C26222T | | | C29738T | | |
| USA/DE-CDC-2-3693752/2020\|EPI_ISL_803105\|2020-10-20 | C11919T | C14408T | T15756G | C15879T | C16293T | C16887T | A20268G | C21214T | A23403G | A23598G | | G25922T | | C28957T | C29642T | C29738T | | |
| USA/DE-CDC-2-3693674/2020\|EPI_ISL_803106\|2020-10-21 | C11919T | C14408T | T15756G | C15879T | C16293T | C16887T | A20268G | C21214T | A23403G | A23598G | | G25922T | | C28957T | C29642T | C29738T | | |

#### B.1.189 S:N679S AA Substitutions

| Sample | ORF10 | | | ORF1a | | | | | ORF1b | | ORF3a | Spike | |
|---|---|---|---|---|---|---|---|---|---|---|---|---|---|
| CNH-B-40 | | ORF1a:K564Q | | | ORF1a:P2018L | | | ORF1a:S3885F | ORF1b:P314L | ORF1b:H2583Y | ORF3a:S177I | S:D614G | S:N679S |
| USA/VA-DCLS-1853/2020\|EPI_ISL_614077\|2020-09-23 | | ORF1a:K564Q | | ORF1a:P1803S | ORF1a:P2018L | ORF1a:A2994V | ORF1a:P3359S | ORF1a:S3885F | ORF1b:P314L | ORF1b:H2583Y | ORF3a:S177I | S:D614G | S:N679S |
| USA/MD-MDH-0280/2020\|EPI_ISL_629060\|2020-10-13 | | ORF1a:K564Q | | | ORF1a:P2018L | ORF1a:A2994V | ORF1a:P3359S | ORF1a:S3885F | ORF1b:P314L | ORF1b:H2583Y | ORF3a:S177I | S:D614G | S:N679S |
| USA/MD-MDH-0281/2020\|EPI_ISL_629061\|2020-10-13 | | ORF1a:K564Q | | | ORF1a:P2018L | ORF1a:A2994V | ORF1a:P3359S | ORF1a:S3885F | ORF1b:P314L | ORF1b:H2583Y | ORF3a:S177I | S:D614G | S:N679S |
| USA/MD-MDH-0282/2020\|EPI_ISL_629062\|2020-10-13 | | ORF1a:K564Q | | | ORF1a:P2018L | ORF1a:A2994V | ORF1a:P3359S | ORF1a:S3885F | ORF1b:P314L | ORF1b:H2583Y | ORF3a:S177I | S:D614G | S:N679S |
| USA/MD-MDH-0353/2020\|EPI_ISL_648292\|2020-10-20 | | ORF1a:K564Q | | | ORF1a:P2018L | ORF1a:A2994V | ORF1a:P3359S | ORF1a:S3885F | ORF1b:P314L | ORF1b:H2583Y | ORF3a:S177I | S:D614G | S:N679S |
| USA/MD-MDH-0354/2020\|EPI_ISL_648293\|2020-10-20 | | ORF1a:K564Q | | | ORF1a:P2018L | ORF1a:A2994V | ORF1a:P3359S | ORF1a:S3885F | ORF1b:P314L | ORF1b:H2583Y | ORF3a:S177I | S:D614G | S:N679S |
| USA/DE-CDC-2-3693752/2020\|EPI_ISL_803105\|2020-10-20 | ORF10:Q29* | ORF1a:K564Q | ORF1a:C1114F | | | | | ORF1a:S3885F | ORF1b:P314L | ORF1b:H2583Y | ORF3a:S177I | S:D614G | S:N679S |
| USA/DE-CDC-2-3693674/2020\|EPI_ISL_803106\|2020-10-21 | ORF10:Q29* | | ORF1a:C1114F | | | | | ORF1a:S3885F | ORF1b:P314L | ORF1b:H2583Y | ORF3a:S177I | S:D614G | S:N679S |
