## Supplemental_files_1_through_4 for "Novel SARS-CoV-2 spike variant identified through viral genome sequencing of the pediatric Washington D.C. COVID-19 outbreak": S4_gisaid_hcov-19_acknowledgement_table_high_quality_spike_n679S.pdf

We gratefully acknowledge the following Authors from the Originating laboratories responsible for obtaining the specimens, as well as the Submitting laboratories where the genome data were generated and shared via GISAID, on which this research is based.

All Submitters of data may be contacted directly via www.gisaid.org

Authors are sorted alphabetically.

| Accession ID | Originating Laboratory | Submitting Laboratory | Authors |
|---|---|---|---|
| EPI_ISL_614077 | Virginia DCLS | Virginia DCLS | Virginia DCLS |
| EPI_ISL_629060, EPI_ISL_629061, EPI_ISL_629062 | Maryland Public Health Laboratory | Maryland Public Health Laboratory | Maryland Department of Health Laboratories Administration |
| EPI_ISL_648292, EPI_ISL_648293 | MD PHL | MD PHL | Maryland Department of Health Laboratories Administration |
| EPI_ISL_688987 | Pathogen Genomics Center, National Institute of Infectious Diseases | Pathogen Genomics Center, National Institute of Infectious Diseases | Tsuyoshi Sekizuka, Kentaro Itokawa, Rina Tanaka, Masanori Hashino, Makoto Kuroda |
| EPI_ISL_693192 | Royal Hobart Hospital | Royal Hobart Hospital | Rob Vanhaeften, Dr Louise Cooley |
| EPI_ISL_693193, EPI_ISL_693194 | Royal Hobart Hospital | Royal Hobart Hospital | Robert Vanhaeften, Dr Louise Cooley |
| EPI_ISL_729818 | Laboratorio Central de Saude Publica do Estado do Rio Grande do Sul (LACEN-RS) | Laboratory of Respiratory Viruses and Measles, Oswaldo Cruz Institute, FIOCRUZ | Paola Resende, Luciana Appolinario, Fernando Motta, Anna Carolina Paixão, Ana Carolina Mendonça, Tatiana Schaffer Gregianini, Marilda Tereza Mar da Rosa, Marilda Siqueira |
| EPI_ISL_803105, EPI_ISL_803106 | Respiratory Viruses Branch, Centers for Disease Control and Prevention | Respiratory Viruses Branch, Centers for Disease Control and Prevention | Queen,K., Li,Y., Tao,Y., Uehara,A., Montmayeur,A., Paden,C.R., Cook,P.W., Marine,R., Sheth,M., Wang,H., Lee,J., Tong,S. |
